# Clustered regularly interspaced short palindromic repeats (CRISPR)-Diagnostics for differential detection of haemoglobin variant S, C, D and E

**DOI:** 10.1101/2025.08.26.25334459

**Authors:** Patrick Adu, Prabhleen Kaur, Prosad Kumar Das, Parikshit Chattopadhyay, C Afzal, Debojyoti Chakraborty

## Abstract

Although haemoglobin variants are prevalent in low- and middle-income countries, the exact disease burden remains unknown due to a lack of diagnostic capacity. Traditionally, routine clinical haemoglobin variant diagnostics have relied on electrophoresis, which separates the haemoglobins based on size and charge differences. However, electrophoresis-based assays are limited in depth and coverage due to their inability to separate co-migrating variants at the pH employed. Importantly, for genetic counselling of pre-marital couples and prenatal screening of inherited haemoglobinopathy risk, molecular-based assays are required. Here, we leveraged the specificity and dual mismatch intolerance of en31FnCas9 to achieve differential identification of haemoglobin variants S, C, D, and E. Moreover, we demonstrate reliable differential detection of homozygous, heterozygous and compound states of haemoglobin variants due to en31FnCas9’s intolerance to dual mismatch in the respective gRNA-target DNA complementarity. Furthermore, we coupled our en31FnCas9-based haemoglobin variant detection to the signal enhancement of recombinase polymerase amplification (RPA) to achieve reliable differential detection of haemoglobin variants from non-invasive saliva and urine samples in <60 minutes. Taken together, our study demonstrates the feasibility of functionalization of CRISPR-based diagnostics as a point-of-care technique towards achieving the democratisation of haemoglobin variant diagnosis in resource-limited settings.

## Introduction

Haemoglobinopathies are genetically inherited disorders of haemoglobin that may result in reduced haemoglobin quantities (thalassaemia) or structural haemoglobin types other than what is physiological for an individual’s age (structural variants). With approximately 7% of the global population (>500 million people) estimated to be carrying haemoglobinopathies [1, 2], haemoglobinopathies are a significant public health problem. Although various hypotheses have been proposed as the basis for the disproportionate sub-Saharan Africa (SSA) and South-East Asia (SEA) representation in the haemoglobinopathies prevalence data, the evolutionary-based theory of selective pressures imposed by the *Plasmodium* parasites may provide a plausible explanation for the co-selection of haemoglobinopathies in *Plasmodium falciparum* endemic regions [3]. The presentations of the various haemoglobinopathies are clinically variable. Thalassaemia, a quantitative globin chain disorder, is haematologically characterised by microcytic red blood cells and increased RBC haemolysis due to precipitation of excess beta or alpha globin chains [4]. Contrastingly, structural haemoglobin variants present as normocytic haemolytic anaemia with variable clinical presentation depending on the exact haemoglobin variant inherited [5]. The most common beta haemoglobin variants of clinical significance are S (an exon 1 point mutation, Glutamic acid to Valine substitution, β^6GAG→GTG^), C (an exon 1 point mutation, Glutamic acid to Lysine substitution, β^6GAG→AAG^), D (an exon 3 point mutation, Glutamic acid to Glutamine substitution, β^121GAA→CAA^), and E (an exon 1 point mutation, Glutamic acid to Lysine substitution, β^26GAG→AAG^). It should be noted that since the inheritance of each of these haemoglobin variants is independent of each other, compound inheritance is a real clinical possibility, particularly in endemic areas.

Since these haemoglobinopathies are genetically inherited, clinical diagnosis should ideally be on genotype-based platforms. These genotype-based assays, such as PCR-based assays (e.g. allele-specific PCR, Gap-PCR) and various sequencing platforms, are, however, not routinely available in regions where haemoglobinopathies are prevalent. In SSA and across the majority of Low- and Middle-Income Countries (LMICs), the lack of molecular-based diagnostic capacity across the healthcare ecosystem has necessitated over-reliance on phenotype-based assays such as haemoglobin solubility and cellulose acetate electrophoresis [6, 7]. Although these phenotype-based assays are economical, their inherent limitations are such that resolving compound inherited variants is not always possible. For example, haemoglobin variants S, D and G are known to co-migrate under the alkaline conditions employed in cellulose acetate electrophoresis, just as haemoglobin variants C, E and O also co-migrate [7, 8]. Thus, in the absence of additional diagnostic assays, underdiagnosis of patients in resource-limited settings is a practical clinical challenge. To overcome this limitation of the routinely employed cellulose acetate electrophoresis, some healthcare facilities combine it with acid electrophoresis, which not only increases the sample-to-result time but also increases the cost for patients. Efforts to expand accessibility to affordable diagnostics have seen the deployment of many point-of-care test devices like the Gazelle, HemoTypeSC, and Sickle Scan; however, none of these overcome the inherent limitations of the clinically widely used cellulose acetate electrophoresis method [9, 10]. Given the diagnostic challenges, it is nearly impossible to estimate the epidemiology and/or disease burden of haemoglobinopathies in LMICs. Consequently, the absence of such concrete data also means that estimating the economic contribution of haemoglobinopathies to the healthcare system in LMICs to aid planning is not an easy task.

CRISPR-based diagnostic platforms (CRISPR-Dx) offer unique opportunities for expanding molecular-based diagnostic access to resource-limited settings. Given the specificity of the guide RNA (gRNA) to its target nucleic acid sequences, CRISPR-based assays can be used to enable differential detection of haemoglobin variants in heterozygous, single homozygous, or compound inherited haemoglobin variant states. Notably, >1000 point mutations [5, 11] have been identified as leading to a variety of haemoglobin variants, creating a clinical scenario that necessitates diagnostic platforms with functionalized multiplexing potential for exhaustive detection. Previously, our laboratory demonstrated that FnCas9, based on its sensitivity to mismatch intolerance [12], can enable the detection of sickle haemoglobin (Hb S) in the FELUDA assay [13]. Here, we demonstrate how the engineered variant-31 of FnCas9 (en31FnCas9) [14] can differentially detect haemoglobin variants S, C, D and E as well as wild-type sequences of the respective regions of these variants. Additionally, we demonstrate the feasibility of coupling haemoglobin variant detection to recombinase polymerase amplification (RPA) to minimise the test turnaround time to facilitate the potential deployment as a point-of-care system. Together, we anticipate that further optimisation of our reported haemoglobin variant detection assay could lead to a robust, sensitive and specific molecular-based assay that can be implemented as a POCT to enable population-based screening in a proactive approach towards reducing the penetrance of haemoglobinopathies.

## Materials and methods

### Study design

The schematic of the relative positions of the haemoglobin variants in the specific exon of the beta globin chain is illustrated in Figure 1. The primer sequences used for the study reported herein are shown in Table 1.

**Figure 1:**
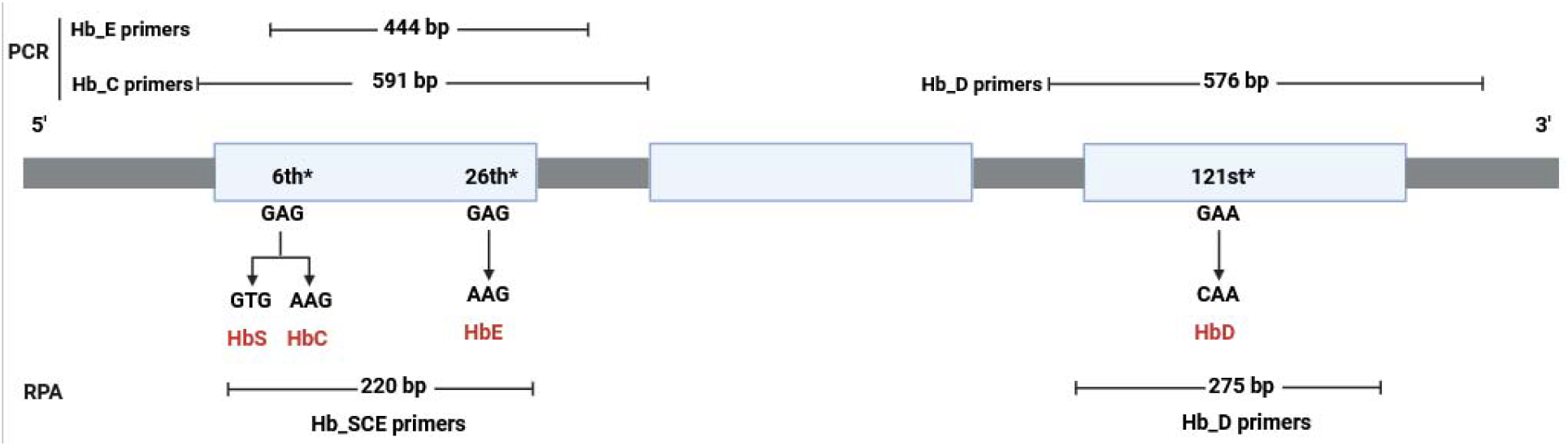
A schematic of the relative positions of the haemoglobin S, C, D, and E variants in the beta globin chain. The Figure is not drawn to scale. (RPA: Recombinase Polymerase Amplification; PCR: Polymerase Chain Reaction; *indicates the codon within which the respective haemoglobin variant point mutation occurs). The figure was created in https://BioRender.com.

**Table 1:**
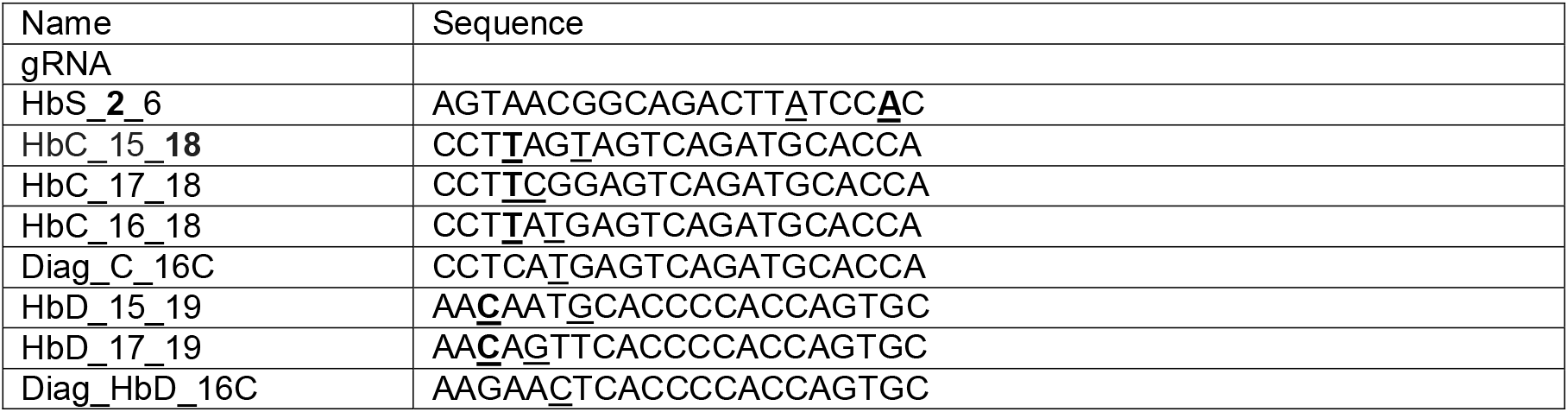
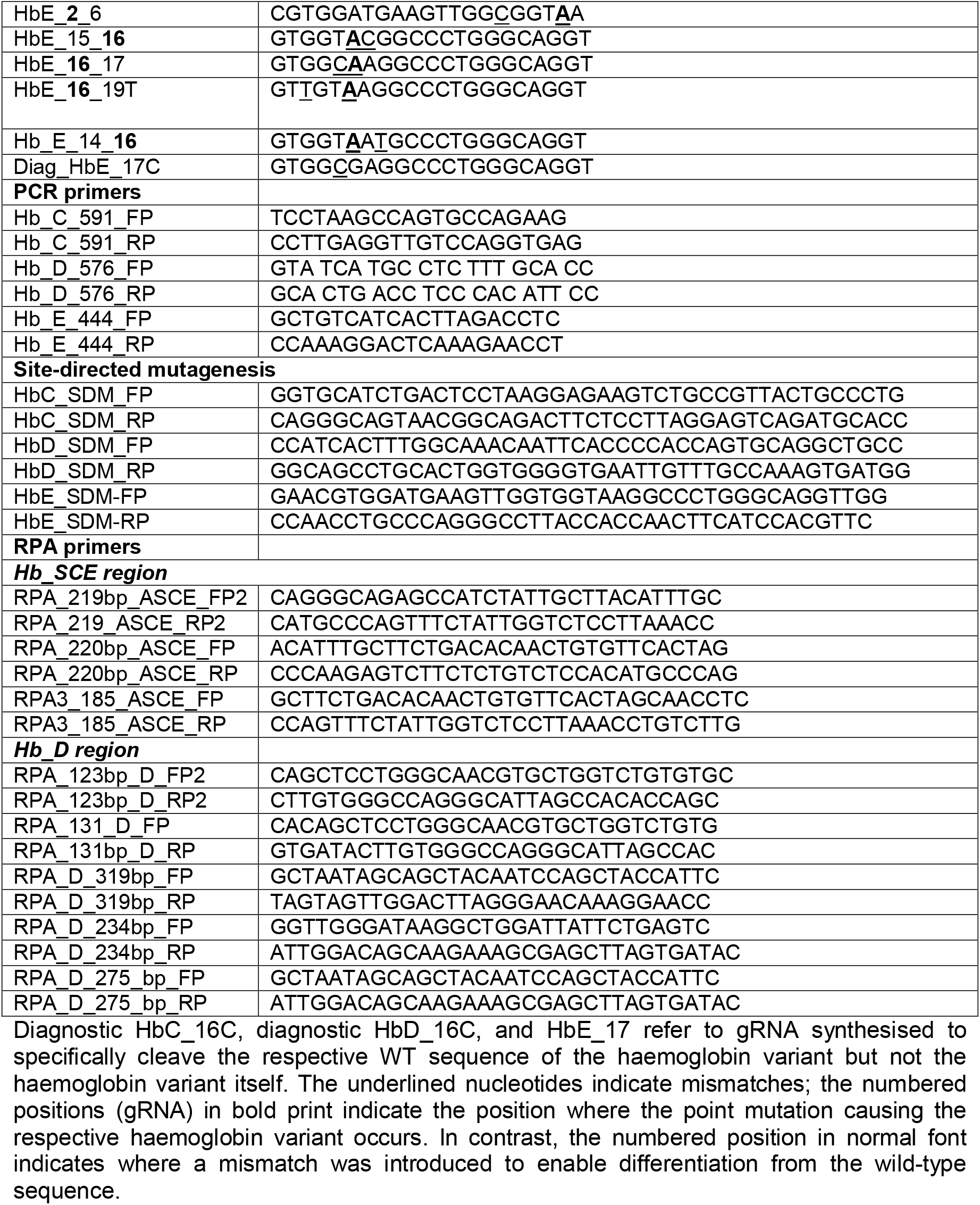
Oligonucleotides used in the present study.

Diagnostic HbC_16C, diagnostic HbD_16C, and HbE_17 refer to gRNA synthesised to specifically cleave the respective WT sequence of the haemoglobin variant but not the haemoglobin variant itself. The underlined nucleotides indicate mismatches; the numbered positions (gRNA) in bold print indicate the position where the point mutation causing the respective haemoglobin variant occurs. In contrast, the numbered position in normal font indicates where a mismatch was introduced to enable differentiation from the wild-type sequence.

### En31Fncas9 Protein purification

The proteins used in this study were purified as reported previously [12]. The plasmid for the en31FnCas9 protein was expressed in *Escherichia coli* Rosetta2 (DE3) (Novagen), and cultured at 37°C in LB medium (supplemented with 50 mg/ml kanamycin and 35 mg/ml chloramphenicol) until OD600 reached 0.6. Subsequently, protein expression was induced by the addition of 0.5 mM isopropyl β-D-thiogalactopyranoside (IPTG). The Rosetta2 (DE3) cells were then cultured overnight (at 18 °C) and harvested by centrifugation. After resuspension of the cells in a lysis buffer (20 mM HEPES, pH 7.5, 500 mM KCl, 5% glycerol supplemented with 1× PIC [Roche], 100 μg/mL lysozyme), the *E. coli* cells were lysed by sonication. After centrifugation, the lysate was affinity-purified by Ni-NTA beads (Roche). The eluted en31FnCas9 protein was further purified by Ion exchange chromatography in which we passed the diluted eluate through pre-equilibrated SP Sepharose beads (Cytiva). The concentration of purified protein was measured by a Pierce BCA protein assay kit (Thermo Fisher Scientific), keeping BSA as a standard. The purified proteins were stored at -80 °C until further use. For all *in vitro* cleavage assays, the en31FnCas9 was reconstituted at 5 µM concentrations, and 1 µl was used in a 10 µl IVC reaction to achieve a 500 nM protein concentration.

### Oligonucleotides used in the study

All the oligonucleotides (Table 1) used in this study were synthesised by Eurofins Genomics, India.

### Plasmid Construction

Human Genomic DNA was extracted from the blood using the DNeasy Blood & Tissue kit (Qiagen) as per the manufacturer’s instructions. WT sequences corresponding to 444 bp (for Hb S and E), 591 bp (Hb C), and 576 bp (Hb D) were PCR-amplified and column-purified for subsequent cloning experiments. The column-purified PCR products of the respective WT sequences of the regions where the haemoglobin variant (S, C, D and E) point mutation occurs were cloned into the TOPO-TA vector (Thermo Fisher Scientific) and then used for generating haemoglobin variants through site-directed mutagenesis.

### Site-directed mutagenesis to generate haemoglobin variants S, C, D, and E

Haemoglobin variants S, C, D, and E were generated within the TOPO-TA plasmid backbone (Acharya et al., 2019) by QuickChange II site-directed mutagenesis (SDM) kit (Agilent) following the manufacturer’s protocol. The success of the SDM-generated haemoglobin variants was confirmed through Sanger sequencing.

### Sanger Sequencing

The sequencing reaction was carried out in 10μl volume (containing 0.5μl purified DNA, 0.8μl sequencing reaction mix, 2μl 5X dilution buffer, and 0.6μl forward/ reverse primer) using the Big Dye Terminator v3.1 cycle sequencing kit (ABI, 4337454). The amplification conditions were: 3 min denaturation at 95°C, followed by 40 cycles of (10 secs at 95°C, 10 secs at 55°C, 4 min at 60°C) and 10 min extension. Subsequently, the PCR product was purified by mixing with 12μl of 125 mM EDTA (pH 8.0) and incubating at RT for 5 min. 50μl of absolute ethanol and 2μl of 3M NaOAc (pH 4.8) were then added, incubated for 10 min at RT and centrifuged for 30 min at 3800rpm, followed by an invert spin at <300rpm to discard the supernatant. The pellet was washed twice with 100 μl of 70% ethanol at 4000rpm for 15 min, and the supernatant was discarded by invert spin. The pellet was air-dried, dissolved in 12 μl of Hi-Di formamide (Thermo Fisher, 4311320), denatured at 95°C for 5 min followed by a snap chill, and linked to an ABI 3130xl sequencer. Base-calling was carried out using sequencing analysis software (v5.3.1) (ABI, US), and the sequence was analyzed using Chromas v2.6.5 (Technelysium, Australia).

### NaOH-based genomic DNA extraction for RPA assay

Genomic DNA (gDNA) extraction from human blood, saliva, or urine was performed through a previously published manual NaOH extraction protocol [15]. Briefly, saliva or urine was mixed with an equal volume of 1X PBS and centrifuged at 13000 rpm. For blood samples, an equal volume of RNAse-free water was added to haemolyse the red blood cells, and centrifuged to sediment nucleated cells. After discarding the supernatant, this was followed by three 1X PBS washes. After the final washing, the pellet was lysed with 40µl of 0.2M NaOH and incubated at 70°C for 10 minutes for urine and saliva pellet, (5 minutes for the blood pellet). Subsequently, the reaction was neutralised with 180µl of 0.04 Tris-HCl (pH 7.5), and centrifuged at 13000rpm, after which the supernatant was transferred into a fresh vial. A total volume of 1 – 2 µl supernatant was used in the PCR or RPA reaction or stored at -20 °C until further use.

### Annealing-extension PCR for making sgRNA template

For each gRNA production, DNA template for in vitro transcription (IVT) was produced through overlap extension PCR by assembling the following on ice: 12.5 μl forward primer (10 μM), 12.5 μl reverse primer (10 μM), 5 μl 10X PCR buffer, 4 μl dNTPs (2.5mM), 0.5 μl Taq pol, 15.5 μl nuclease-free water to make a total volume of 50 μl reaction. The forward primer was unique for each haemoglobin variant; the reverse primer was, however, common for all the haemoglobin variants (Table 1). The amplification conditions were: Initial denaturation (95□C, 3min), 35 cycles of 95□C (10s), 55□C (30s), 72□C (30s), followed by 7 minutes’ final extension. The size of the PCR product was verified on a 2% agarose gel.

### In Vitro Transcription (IVT) Reaction

All the gRNAs for haemoglobin variants were 21-nucleotide sequences with a single base mismatch insertion to ensure that although there was only a single mismatch with the haemoglobin variant, there was a dual mismatch with the WT sequence (see Table 1). *In vitro* transcription for gRNAs/crRNAs was done using the MegaScript T7 Transcription kit (ThermoFisher Scientific) following the manufacturer’s protocol. Briefly, IVT was set up in a 20 µl total volume comprising 2μl of 10X Reaction Buffer, 2μl of UTP (75mM), 2μl of ATP (75mM), 2μl of GTP (75mM), 2μl of CTP (75mM), 5μl extension PCR product as template, 2μl T7 polymerase and 3 μl of nuclease-free water as per the manufacturer’s protocol. IVT reactions were incubated at 37°C overnight (>16 hours) for sgRNA synthesis. Following DNase I treatment (Thermo Fisher Scientific) to remove residual DNA, gRNA clean-up was undertaken using Monash RNA purification columns as per the manufacturer’s protocol. The respective gRNA concentration was quantified on a NanoDrop spectrophotometer (Thermo Fisher Scientific). Purified IVT sgRNAs/crRNAs were stored at -20°C until further use. For all *in vitro* cleavage assays, the sgRNA was reconstituted at 5 µM concentrations, and 1 µl was used in a 10 µl IVC reaction to achieve a 500 nM sgRNA concentration (unless otherwise stated).

### Detection via *In Vitro* Cleavage (IVC)

PCR-amplified DNA products of respective haemoglobin variants were used as substrates for *in vitro* cleavage assays as previously published [12]. 9 μl RNP of the en31FnCas9 protein and respective gRNA was reconstituted by combining 10x CutSmart buffer (1μl), 50mM MgCl (2μl), 5 μM en31FnCas9 protein (1μl), 5 μM gRNA (1.1 μl), and 3.9 μl nuclease-free water (at 25°C for 20 minutes). For each IVC assay, 1 μl of purified target DNA amplicon (500 nM to achieve a final 50 nM concentration in a 10 µl reaction volume) was added to the preformed RNP and incubated at 37°C for 5 – 30 minutes, as the case may be; cleaved products were visualised on the 2% agarose gel.

### RPA

For screening of RPA primers, three forward primers and three reverse primers flanking the site for gRNA for Hb S, C, and E regions (for Hb D region, five primer pairs were tested) were designed (Figure 1 for location and Table 1 for primer sequence). Each primer was 30 – 34 bp long, and amplicons resulting from all primer combinations were between 120 and 319 bp long. The TwistAmp® Basic kit was purchased from TwistDx Ltd. (Hertfordshire, AL, UK). Basic Kit (TwistDx) and used for isothermal amplification of gDNA following the manufacturer’s protocol with few modifications. Briefly, RPA reactions were undertaken in a final 25 μL volume comprising 20 μL rehydrated pellet (29.5 μL of primer-free rehydration buffer, one pellet and 9.5 μL nuclease-free water), 1 μl 0.25 μM forward, 1μl 0.25 μM reverse primer, 2 µl of NaOH-extracted gDNA, and 1 μl 280mM magnesium acetate (last added). After vortexing and a brief spin of the mixture, it was incubated at 37 °C for 20 – 30 minutes. Subsequently, the amplification products were either column-purified and resolved on a 2% agarose gel or used directly for en31FnCas9–sgRNA IVC reaction.

## Results

### Differential detection of haemoglobin S, and C variants from wild-type sequence

The point mutations leading to haemoglobin S (β^6GAG→GTG^) and C (β^6GAG→AAG^) variants occur in the sixth codon of the beta globin chain. Thus, differential detection of these haemoglobin variants requires a highly specific assay, given that there is only one nucleotide difference between Hb S and C. Here, we show that when using PAM distal gRNA for Hb S variant that has a single mismatch introduced at position 6 [12], en31FnCas9 enables *in vitro* differential detection of Hb S from wild-type haemoglobin sequence (Figure 2A & B). In Figure 1B, we mimicked the heterozygous Hb AS inheritance by combining the Hb S and WT sequence of the Hb S region in a single tube and then explored whether using a combinatorial gRNA for Hb S and WT could aid differential detections of each of the amplicons. The specificities of the gRNA enabled distinct identification of heterozygous Hb AS, indicating that these gRNAs do not interfere with each other when used simultaneously in a single IVC reaction (Figure 2B). Furthermore, under our assay conditions, detection of Hb variants could be visually seen on an agarose gel from 10 minutes of 37°C incubation (Figure 2A). Moreover, when using PAM distal gRNA specific to Hb C variant with a single mismatch introduced at the 15, or 16, or 17^th^ position, the en31FnCas9 enabled differential detection of Hb C variant (Figure 2C). Furthermore, we also demonstrate that the gRNA used for these Hb variant detections did not show cleavage towards the wild-type sequence of the same region (Figure 2C).

**Figure 2:**
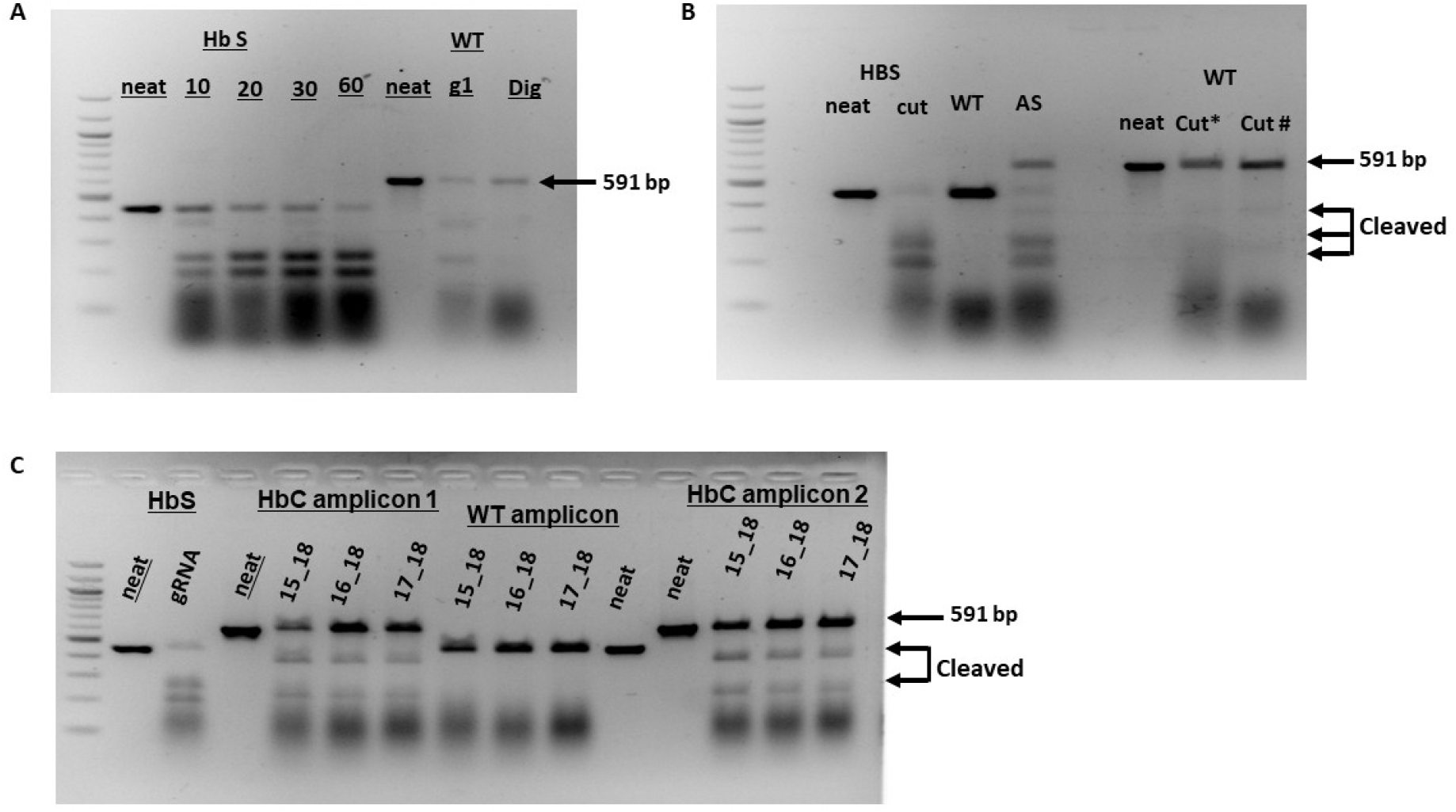
gRNA for differential detection of Hb S, C and WT sequences. A depicts a time-course IVC for Hb S amplicon (using Hb S gRNA) and WT amplicon (using WT-specific gRNA for the WT sequence of the Hb S region, and Diag_gRNA able to detect heterozygous inheritance). B is IVC for the differential detection of homozygous S and heterozygous S (AS). C is the optimisation of gRNA for the differential detection of homozygous Hb C inheritance. [15_**18**, 16_**18**, & 17_**18** identify positions relative to the NGG PAM where mismatches were introduced to enable differential detection of Hb C variant]. Note that the numbered positions in bold print indicate the position where the point mutation causing the respective haemoglobin variant occurs. In contrast, the numbered position in normal font indicates where a mismatch was introduced to enable differentiation from the wild-type sequence.

### En31FnCas9 enables differential detection of Hb D and E variants in *in vitro* IVC

Haemoglobin D point mutation occurs in exon 3 of the beta globin chain (β^121GAA→CAA^). When using PAM distal gRNA specific to the haemoglobin D variant with a mismatch introduced at the 15^th^ or 17^th^ position, we demonstrate that en31FnCas9 enabled differential detection of the Hb D variant (Figure 3A). Additionally, neither the 15_19 gRNA nor the 17_19 gRNA cleaved the wild-type sequence of the Hb D region. In Figure 3B, we demonstrate that both PAM-proximal and PAM-distal gRNA can enable the differential detection of haemoglobin variant E. The respective PAM proximal (HbE 2_6 gRNA) and distal (HbE 16_19 gRNA) guides did not show activity against the wild-type sequence of the HbE region, showing that these gRNAs can be employed to differentiate the homozygous state from a wild-type sequence. Since the haemoglobinopathy genes are recessively inherited, identification of both heterozygous and homozygous states is important for genetic counselling. In the present study, we demonstrate that when a single mismatch is introduced into the WT gRNA, in a position other than where the haemoglobin variant point mutation occurs, ensuring that the gRNA has two mismatches against the respective haemoglobin variant, en31FnCas9 can be used to enable differentiation of the WT sequences from the haemoglobin variants (Figure 3C). In leveraging the intolerance of en31FnCas9 to dual mismatches, we demonstrate that WT sequences of the haemoglobin variants C, D, and E can be differentially identified from their respective variants using diagnostic HbC_16C, diagnostic HbD_16C, and HbE_17, respectively (Figure 3C). Multiplexing functionalization represents an important step in achieving point-of-care test democratisation as it improves test throughput and assay affordability through the reduction of the cost per test. A crucial step in navigating the multiplexing potential of the CRISPR-Dx is ascertaining the non-interference of the functionalities and specificities of gRNAs. In this regard, we mimicked compound inherited haemoglobinopathies by combining haemoglobin variants in a single tube, and then explored the feasibility of combinatorial gRNAs to simultaneously detect these through in vitro cleavage assays (Figure 3D). Our IVC data demonstrate that potentially inherited compound haemoglobinopathies CE and DE can be reliably differentially detected using en31FnCas9 (Figure 3D).

**Figure 3:**
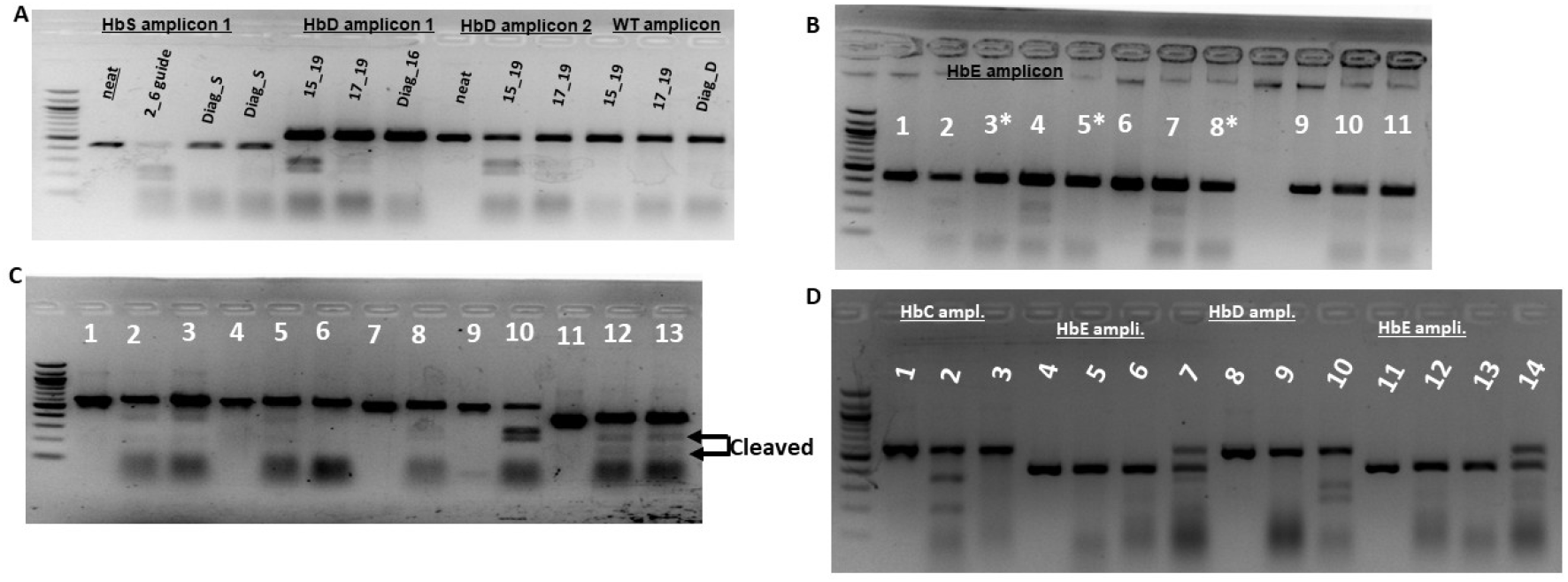
gRNA for differential detection of Hb C, D, E and WT sequences. A is the optimisation of gRNA for the differential detection of homozygous Hb D inheritance. [15_**19**, & 17_**19** identify positions relative to the NGG PAM where mutations were introduced to enable differential detection of Hb D variant]. **Figure B** is differential detection of Hb E amplicon; [lanes 1, 6 & 9 are uncut; *indicates that lane (3, 5 & 8) were cleaved with gRNA specific to WT sequence of the Hb E region; lanes 2 (gRNA_**2**_6), 4 (gRNA_**16**_19) and 7 (gRNA_14_**16**) were cleaved with Hb E specific gRNA; lane 9 is WT sequence of Hb E region uncut; lanes 10 and 11 are WT sequence of HbE region cleaved with Diag_HbE_16 and Diag_HbE_17 respectively. **Figure C** is the differential detections of Hb C, D and E (Lane 1: HbC amplicon uncut, Lane 2: HbC cleaved with HbCgRNA_16_**18**, Lane 3: HbC cleaved with HbCgRNA_17_**18**, Lane 4: WT sequence of HbC region uncut, Lane 5: WT sequence of HbC region cleaved with Diag_HbC_16C, Lane 6: WT sequence of HbC region cleaved with Diag_HbC_17C, Lane 7: HbD uncut, Lane 8: HbD cleaved with Diag_HbD_16C, Lane 9: WT sequence of HbD region uncut, Lane 10: WT sequence of HbD region cleaved with Diag_HbD_16C, Lane 11: HbE uncut, Lane 12: HbE cleaved with, HbE_14_**16**_gRNA; Lane 13: WT sequence of HbE region cleaved with Diag_HbE_17. **Figure D** shows that the combinatorial use of gRNA with specificities for different haemoglobin variants does not interfere with each other: Lane 1: HbC uncut, lane 2: HbC cleaved with HbC_15_**18** gRNA, Lane 3: HbC cleaved with HbE_14_**16** gRNA, lane 4: HbE uncut, lane 5: HbE cleaved with HbC_15_**18** gRNA, lane 6: HbE cleaved with HbE_14_**16** gRNA, Lane 7: Combined HbC & HbE variants cleaved with HbC_15_**18** gRNA and HbE_14_**16** gRNA, lane 8: HbD uncut, lane 9: HbD cleaved with HbE_14_**16** gRNA, lane 10: HbD cleaved with HbD_15_**19** gRNA, lane 11: HbE uncut, lane 12: HbE cleaved with HbE_14_**16** gRNA, lane 13: HbE cleaved with HbD_15_**19** gRNA, lane 14: combined HbD and E variants cleaved with HbD_15_**19** gRNA and HbE_14_**16** gRNA. [Note that the numbered positions in bold print indicate the position where the point mutation causing the respective haemoglobin variant occurs. In contrast, the numbered position in normal font indicates where a mismatch was introduced to enable differentiation from the wild-type sequence.]

### RPA-based haemoglobin variant detection from less invasive urine and saliva samples

Since there is no tool to predict the efficiency of RPA primers, we screened a series of primers for isothermal amplification of the genomic regions containing the haemoglobin S, C, E variants on one hand, and haemoglobin D on the other hand (See table 1). We initially optimised the NaOH gDNA extraction procedure [15] using 30 ml of urine donated by two individuals. As shown in Figure 4A, 1 – 2 µl of the NaOH-extracted gDNA was successfully amplified using the traditional PCR method. We were further able to successfully amplify gDNA extracted from 7 ml or 15 ml urine using the NaOH extraction method (Figure 4B). Subsequently, we were able to achieve reliable PCR amplification from as little as 500 µl saliva samples (Figure 4C). Among the many limitations of the traditional PCR-based assay for point-of-care diagnostics are the long processing times and the requirement for thermal cyclers, which are not routinely available in LMICs. Thus, we explored RPA assay for the detection of the haemoglobin variants. In Figure 4D and E, we demonstrate the feasibility of successfully implementing RPA to amplify NaOH-extracted gDNA from blood, urine and saliva. Furthermore, we demonstrate that 1 – 2 µl of the RPA product directly used in an *in vitro* cleavage assay enabled the detection of haemoglobin variant within 50 – 60 minutes (10 - 15, 20, and 20 minutes for NaOH extraction, RPA and 20 minutes for IVC, respectively). Taken together, our data demonstrate the potential of implementing a molecular-based isothermal haemoglobin variant detection assay, which is suitable for resource-limited settings.

**Figure 4:**
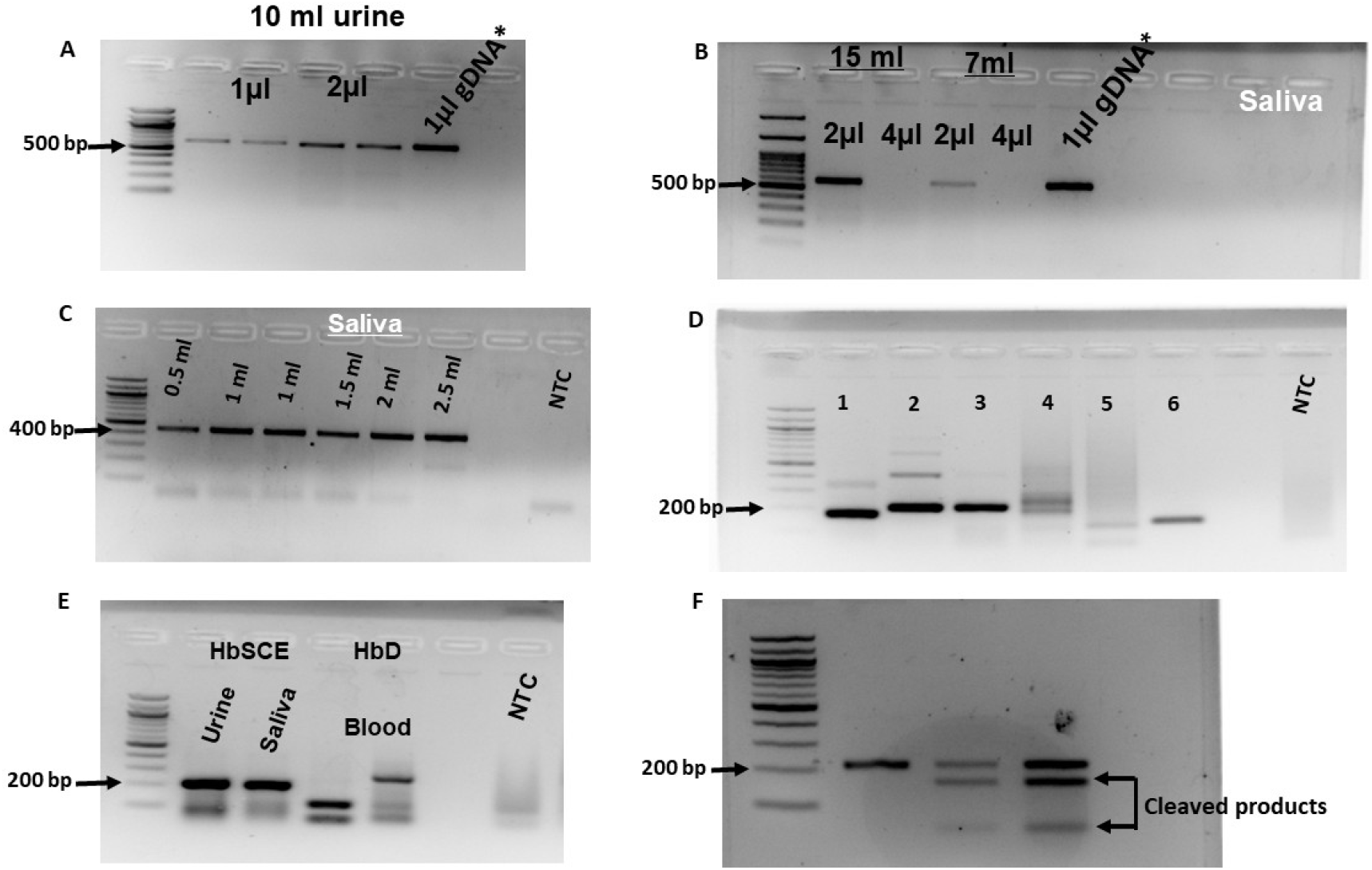
Optimisation of less invasive samples for Recombinase Polymerase Amplification assay. A is a 10 ml of urine volume from two donors (amplified PCR products are 591 bp) used for NaOH-based gDNA extraction; B shows gDNA 7 and 15 ml saliva from two donors (amplified PCR products are 591 bp) used for NaOH-based gDNA extraction; C shows the titration of the minimal saliva volume that can be used for NaOH-based gDNA extraction for PCR amplification (amplified PCR products are 444 bp); D is optimisation of primers for RPA for the amplification of Hb SCE and Hb D regions (amplified RPA products are 190 bp, and 220 bp for HbSCE primers); E is RPA for Hb SCE region and HbD region using NaOH-based gDNA extracted from urine, saliva and blood (amplified PCR products are 220 bp for HbSCE region and 131 bp and 275 bp for HbD region); F is an in vitro cleavage assay undertaken on 1 µl or 2 µl volume of RPA amplified Hb C variant. (*indicates 1µl of Qiagen column purified gDNA from blood)

## Discussion

Clinically, haemoglobin variant diagnosis in LMICs has traditionally relied on phenotype-based assays such as cellulose acetate and/or acid electrophoresis, and solubility assays. While these assay platforms are economically feasible in resource-limited settings, their inherent failure to resolve co-migrating haemoglobin bands may inadvertently lead to underdiagnosis in some cases [7, 8]. The inherent sensitivity, specificity and scalability of CRISPR-based diagnostics offer unique opportunities for achieving democratisation of haemoglobinopathy diagnostics. Here, we leverage the intolerance of en31FnCas9 to dual mismatch in the PAM proximal or distal regions of the gRNA to demonstrate differential detection of haemoglobin variants S, C, D and E. Furthermore, we demonstrate that the inherent specificity of the en31FnCas9 can also be leveraged to achieve differentiation of the homozygous and heterozygous states.

There are hundreds of genetically inherited point mutations that lead to haemoglobin variants and, in some cases, thalassaemia. Given that each of these point mutations is independently inherited, compound inherited mutations causing complex clinical presentations or symptomatology are likely scenarios in areas with high haemoglobinopathy burden. Thus, diagnostic platforms with multiplexing functionality have the potential to expand the diagnostic spectrum for these haemoglobinopathies in LMICs. Deploying such multiplex diagnostics in a POCT format in LMICs could potentially increase accessibility to neonatal screening and premarital screening programs to improve healthcare delivery as well as empower individuals to make informed choices to reduce the haemoglobinopathy burden. CRISPR diagnostics, as a molecular-based assay, has the potential to meet these key metrics given its nucleic acid-based specificity. Although our study focused on a limited number of haemoglobin variants of clinical significance, the differential *in vitro* detections achieved through the combination of gRNAs, even in heterozygous states, attest to the untapped potential of CRISPR-based diagnostics in multiplexed detection of compound haemoglobin variants. One of the key hindrances to the deployment of molecular-based assays in resource-poor settings has been the requirement for thermal cyclers to enable biomarker amplification to enhance signal detection. However, the discovery of isothermal amplification methods such as helicase-dependent amplification (HDA) [16], loop-mediated isothermal amplification (LAMP) [17], and recombinase polymerase amplification (RPA) [18] has not only enabled signal amplification without the need for thermal cyclers but also achieved sample-to-result in a relatively shorter time frame. These isothermal-based CRISPR-diagnostics have been previously harnessed for ultra-sensitive identification and diagnosis of various infectious agents [19-21]. In our present study, we demonstrate the compatibility of the haemoglobin variant detection assay with RPA-mediated signal amplification, strengthening the case for its potential point-of-care application. In the clinical settings, haemoglobin variant detection assays have traditionally required blood as a source material [7]; the expertise for phlebotomy limits the performance of these assays to hospital settings. In the present study, we have demonstrated the reliability of using non-invasive samples, such as urine and saliva, as blood substitutes and thereby minimise the requirement for technical expertise when procuring samples. Thus, we anticipate that future protocol optimisation studies, and eventual coupling to a sensitive readout, will enable its deployment in primary healthcare ecosystems in resource-limited settings due to the convenience of saliva or urine sampling.

Except for Hb S, in which we had patient samples, our study was carried out using laboratory-generated site-directed mutagenesis-derived haemoglobin variants and may not account for the nuances associated with patients’ chromosomal background. Thus, future field validation studies involving the use of samples from individuals with these inherited variants are expected to foster better assay characterisation and eventual POCT functionalisation. Notably, since Hb E mutation is associated with thalassaemia symptoms and clinical sequelae [22], our study demonstrates the potential for CRISPR-Dx as a diagnostic tool for point mutation-induced thalassaemia syndromes [23]. Thus, future functionalisation of our molecular-based CRISPR-Dx assay can complement existing NBS and population screening programs in LMICs. Previously, others have demonstrated the potential for cost-saving through the miniaturisation of CRISPR-diagnostics when deployed on a microfluidics platform to improve its sensitivity [24]. Santhiago and colleagues also demonstrated that the sensitivity of CRISPR-Dx was heavily dependent on enzyme kinetics and detector sensitivity [25]. Thus, we anticipate that further characterisation of our reported haemoglobin variant detection assay using various CRISPR-Cas orthologues and exploration of various detection platforms will lead to improved assay read-out options to facilitate the democratisation of haemoglobin variant diagnostics. Although mandatory newborn screening programs have improved patient care in developed economies, such programs have not been consistently implemented in LMICs due to cost implications. Thus, investments in research and development of molecular-based assays such as those described in the present study towards potential POCT functionalisation hold promise for achieving affordable and accessible universal newborn haemoglobinopathy screening.

## Data Availability

All data produced in the present work are contained in the manuscript.

## Declarations

### Data availability

The sequences of the oligonucleotides used in this study are enclosed in Table 1 of the manuscript. All the gel images on which the report is based are also enclosed in the manuscript.

### Research ethics

Our study was approved by the Human Ethics Committees of the CSIR – Institute of Genomics and Integrative Biology, New Delhi. The study was performed in accordance with the principles of the 1975 Helsinki Declaration as revised in 2024. All participants who donated urine, blood and/or saliva samples gave written informed consent before sampling; data were de-identified to preserve participants’ anonymity.

### Consent for publication

All the authors and their respective affiliated institutions gave consent for publication.

### Conflict of interest

D.C. is listed as co-inventor for a US Patent titled, “Kinetically enhanced engineered FnCas9 and its uses thereof” which includes the engineered 31 FnCas9 variant used in this study. The patent applicant (application number is 18/049,291) is the Council of Scientific and Industrial Research, New Delhi, India, and has been granted. The authors have no other competing interests to declare.

## Acknowledgements

P.A. is supported by a CSIR-UNESCO-TWAS Postdoctoral Research Fellowship; P.K., PKD, PC and AC are supported by funding from the Council for Scientific and Industrial Research, India.

## Contributions

P.A. and D.C. conceived the project and designed the experimental protocol. P.C. and A.C. designed and performed the protein engineering experiments. P.A. and P.K. optimised the cloning, site-directed mutagenesis, and IVC assays. P.A., P.K. and P.K.D. designed and implemented bioinformatics tools for the design of gRNAs, validation, and in vitro cleavage assays. P.K. and P.A. were involved in the optimisation of RPA assays. P.A. wrote the draft manuscript. All authors critically reviewed the manuscript. All authors read and approved the final manuscript.

## Notes

### Author Declarations

Our study was approved by the Human Ethics Committee of the CSIR Institute of Genomics and Integrative Biology New Delhi. The study was performed in accordance with the principles of the 1975 Helsinki Declaration as revised in 2024. All participants who donated urine, blood or saliva samples gave written informed consent before sampling; data were de-identified to preserve participants anonymity.

